# Are disturbances in mentalization ability similar between schizophrenic patients and borderline personality disorder patients? A pilot study

**DOI:** 10.1101/2021.11.24.21266797

**Authors:** Tair Bar, Issam Ikshaibon, Muhammad Abu-Alhiga, Tamar Peleg, Yaseen Awad, Eilam Plazur, Idit Golani, Ido Peleg, Alon Shamir

## Abstract

There is a growing interest in theory of mind (ToM) performance among individuals with psychiatric disorders, however, the difference and the performance level between different diagnoses is unclear. Here, we compared the ToM abilities of schizophrenia, schizoaffective and borderline personality individuals (BPD) with healthy individuals, and searched for a correlation between ToM ability, social skills, and empathy. Overall, diagnostic groups performed worse in the Reading the Mind in the Eyes test and recognized fewer Faux Pax motifs than healthy individuals recognize. No difference was observed in the ability to perform the eyes test between schizophrenic patients with and without the affective components and BPD patients. Both schizophrenia and BPD patients received a higher score in the autism-spectrum questioner, while all the diagnostic groups scored lower on the empathy quotient scale than healthy individuals. Finally, a correlation was found between ToM ability and empathy, but not with autistic-related traits. Results indicate that both cognitive and affective aspects of ToM are impaired across all the diagnostic groups, challenge the ability to use ToM as a differential diagnostic tool, and strengthens the correlation between decreased empathy and impaired ToM.

## 1. Introduction

Schizophrenia is associated with a deficit in social functioning and social cognition. Individuals with schizophrenia exhibit substantial and persistent impairments in identifying emotions, social perception, and emotion processing (Green et al., 2015). One of the key components in the functional impairment of social cognition in schizophrenia is the ability to attribute the mental states such as thoughts, beliefs, and intentions of another person. In general, individuals with schizophrenia performed worse in different tasks that assay the individual ability to evaluate and judge the mental state of another (Dodell-Feder et al., 2014; Hegde et al., 2021; Vucurovic et al., 2020). For example, two meta-analyses reported a large magnitude of deficits in understanding that another person has beliefs different from one’s own, and in decoding other’s complex mental states (Emre Bora et al., 2009; Sprong et al., 2007). Besides, individuals with schizophrenia showed deficits in recognition of social Faux Pas, (Negrão et al., 2016). We recently demonstrated that individuals with schizophrenia performed worse in the “Reading the Mind in the Eyes” (eyes) test, a simple well-defined task to infer the mental state of others than healthy individuals (Tadmor et al., 2016). Others and we further suggested that understanding another person’s mental state is a trait marker of the disorder and not dependent on the current state of the illness. We, also, demonstrated that schizoaffective individuals might differ in their ability to decode the mental state of other people from people with schizophrenia and first-episode individuals (Tadmor et al., 2016). However, the question of whether schizophrenia and schizoaffective individuals differ in their ability in decoding the mental state of others and other social cognitive is still open.

Studies examining mentalization have also been conducted among other mental health disorders diagnostic groups (E Bora et al., 2009), including borderline personality disorder (BPD). Borderline personality disorder (BPD) is a debilitating psychiatric condition present in about 1-2% of the general population, 6% of primary care patients, and up to 20% of patients in psychiatric hospitals and outpatient clinics. A meta-analysis found that BPD underperformed in mental state reasoning, and cognitive ToM, but not in decoding the mental state of others (Németh et al., 2018). On the other hand, a recent study found that PBD tends to hypermentalize when they misinterpret social information (Normann-Eide et al., 2020). However, the similarity and the difference in the level of the performance between the different diagnostic groups is less studied.

The relationship between decoding the mental state of others and autistic symptoms had been suggested by Baron-Cohen. Baron-Cohen and his colleagues demonstrate that performance in the eyes test was inversely correlated with performance on the Autism Spectrum Quotient (AQ), a self-report questionnaire that estimates the degree of autistic-related traits in healthy and non-healthy individuals (S Baron-Cohen et al., 2001; Golan et al., 2006). In another word, a high score in the AQ questionnaire (more autistic traits) will probably correlate with impairment performance in the eyes test. Indeed, individuals with autism who characterized by impaired social and communicative functioning displayed poor performance on the eyes task. Furthermore, an eyes test-AQ correlation was found in females, but not males, with autism. On the other hand, inverse eyes test-AQ correlation was found only in healthy men, but not in women (Baron-Cohen et al., 2015). However, it is unknown whether this eyes test-AQ correlation occurs in other psychiatric disorders that associate with social withdrawal, communication impairment, and social cognition deficit.

Here, we compared the mentalization abilities of patients diagnosed with schizophrenia with and without affective component, patients with borderline personality disorder (BPD), with healthy individuals, focusing on both the affective and cognitive components of ToM. Besides, we searched for a correlation between ToM ability, autistic-related traits, and empathy.

## 2. Materials and Methods

### 2.1 Subjects

The Manor Mental Health Center Helsinki committee approved the study (01-19-MZR), and all participants gave informed consent to take part in the study. Forty-four clinically stable individuals with schizophrenia (SZ), 11 individuals with schizoaffective (SZ-AF), and 11 individuals with borderline personality disorder (BPD) meeting the DSM-5 criteria were recruited from the open and closed wards of Mazor Mental Health Center, Akko, Israel. Inclusion criteria were: 1) Males and females 18-60 years of age. 2) Sufficient knowledge of the Hebrew language. The exclusion criteria were: 1) Drug or alcohol abuse, 2) Mental retardation, and 3) Organic brain pathology. Clinical and sociodemographic data were collected from the participant’s electronic medical records and included age, sex, education, ethnicity, age of onset, number of hospitalizations, duration of the illness, and family history. Eighteen healthy individuals without psychiatric history and drug or alcohol abuse served as controls. Sociodemographic information was collected from a self-reported questionnaire and included age, sex, education, and ethnicity.

### 2.2 Reading the Mind in the Eyes test

The “Reading the Mind in the Eyes” (eyes test) test was developed by Baron-Cohen (S Baron-Cohen et al., 2001) as a tool to evaluate the ability to infer the mental state of another person. In this task, participants are presented with 36 still pictures of the eye region of faces illustrating emotionally charged or neutral mental states. They were then asked to choose which of the four words best described what the person in the picture was thinking or feeling. This task is considered an advanced ToM test since the participants need to imagine themselves in the mind of the person shown in the picture. One limitation of the test is that the participants only decode the relevant mental state without predicting or explaining the action of the other person. The score on the eyes test is calculated as the total number of correctly identified mental states.

### 2.3 Faux Pas recognition test

“Faux Pas”: is a term that originated in French, and is used to describe words or behaviors that are not socially acceptable or impolite. The test is based on Gregory et al (Gregory et al., 2002). In brief, the test includes 20 short stories in random order; 10 Faux Pas stories, and 10 stories without Faux Pas motive served as control. The stories have been read out loud, and the subject was asked six questions to identify the “Faux Pas” motif and two-story comprehension questions. For each story containing a Faux Pas, the subject gets 1 point for each question answered correctly. If the subject answer “no” to the first question “if anyone said something that they should not have said”, he or she will get 0 points for that whole story. Overall, a total of 60 points can be scored on the Faux Pas stories. The subject also scores 2 points if he or she recognized that the story did not include a Faux Pas motive (A total of 20 points can be scored on the stories without Faux Pas). In addition, the subject gets 1 point for each story comprehension question answered correctly (overall, a total of 40 points).

### 2.3 The empathy quotient scale (EQ scale)

EQ scale was used to study the subject’s ability to empathize. This questionnaire is intended to measure how easily the subject notices other people’s feelings and how strongly the subject is affected by other people’s feelings. It includes 40 questions related to empathy and 20 control questions. For each of the statements, the subject must choose one from the options: “Totally agree”, “Some agree”, “Some disagree”, or “Do not agree at all”. The average scores on this test are 47 for women and 42 for men (Baron-Cohen and Wheelwright, 2004).

### 2.4 Autism spectrum Quotient (AQ)

The questionnaire consists of 50 sentences in random order related to autism traits (social skills, routine, switching, imagination, and numbers/patterns). In brief, participants are asked to indicate whether they ‘strongly agree’, ‘slightly agree’, ‘slightly disagree’ or ‘strongly disagree’ with each statement. Each item scores zero or one. One point is scored if the participant chooses the ‘autistic trait’ response. A total score is calculated by summing across items (Simon Baron-Cohen et al., 2001; Wheelwright et al., 2010).

### 2.5 Statistical Analysis

Statistical analysis was performed with IBM SPSS statistics 21 and GraphPad Prism 8.2. A normal distribution (Kolmogorov-Smirnova and Shapiro-Wilk tests) and approved homogeneity were analyzed. For normally distributed data, differences among multiple groups were assessed by one-way and two-way ANOVA. Post hoc differences were determined by Tukey’s test and independent-sample t-test when significant main effects or interactions were detected. For non-parametric tests, the Kruskal-Wallis test was conducted when the data were non-normally distributed, followed by Bonferroni correction test. Pearson correlation coefficient was performed to test correlations between variables. Data are presented as mean ± SEM or mean rank. The accepted value of significance for all tests was set at p < 0.05.

## 3. Results

### 3.1 Socio-demographic and clinical characterization

Sixty-six participants were recruited from the open and closed wards of Mazor Mental Health Center for this study, while eighteen healthy individuals served as control. Table 1 presents the demographic and clinical characteristics of the study population. Overall, 62% of the study participants were men. Individuals with schizophrenia with and without the affective component were significantly older (One way ANOVA, F_(3,83)_=3.189, p<0.05) compared with the control and individuals with BPD (Tukey’s, p<0.05), and had fewer years of education (One way ANOVA, F_(3,83)_=31.032, p<0.001) than the control group (Tukey’s, p<0.001). No statistical difference in the number of years of education between BPD and the schizophrenic diagnosis groups was found (Tukey’s, p>0.05). At the clinical levels, no statistically significant differences were found between the study groups in the mean age of the onset and number of hospitalization (One way ANOVA, F_(2,65)_=0.30, p>0.05; Kruskal-Wallis test, χ^2^_(2)_ =2.987, p>0.05, N=66, respectively). However, the duration of the illness was significantly higher in individuals with schizophrenia and schizoaffective than in BPD (One way ANOVA, F_(3,83)_=4.133, p<0.05; Tukey’s, p<0.05).

**Table 1:** Demographic and clinical data by group

|  | **Controls**<br>**(n=18)** | **Schizophrenia**<br>**(n=44)** | **Schizoaff**<br>**(n=11)** | **BPD**<br>**(n=11)** | **p-value** |
| --- | --- | --- | --- | --- | --- |
| **Age (years)** | 34.3±2.8 | 41.2±1.8 | 41.2±3.4 | 30.8±3.1 | **p<0.05<sup>a</sup>** |
| **Sex** |  |  |  |  |  |
| **Men** | 7 (39%) | 36 (82%) | 7 (64%) | 2 (18%) |  |
| **Women** | 11 (61%) | 8 (18%) | 4 (36%) | 9 (82%) |  |
| **Education**<br>**(years)** | 17.1±0.6 | 12.1±0.3 | 12.4±0.4 | 11.2±0.5 | **p<0.001<sup>a</sup>** |
| **Onset (years)** |  | 29.4±1.5 | 28.9±2.8 | 26.7±3.4 | p>0.05<sup>a</sup> |
| **No. of**<br>**hospitalizations** |  | 11.5±1.7 | 9.4±2.6 | 6.7±2.5 | p>0.05<sup>b</sup> |
| **Years of illness** |  | 11.0±1.2 | 12.5±1.9 | 4.7±1.3 | **p<0.05<sup>a</sup>** |

Data presented as a mean ± standard error of the mean (SEM). <sup>a</sup>One-way ANOVA, <sup>b</sup>Kruskal-Wallis test. Abbreviations: Schizoaff= Schizoaffective; BPD= Borderline Personality Disorder

### 3.2 Reading the mind in the eyes is impaired in individuals with schizophrenia, schizoaffective, and BPD

The ability to decode the mental state of another person was evaluated using the eyes test. No statistically significant main effect of sex or interaction between sex and the diagnosis group was found (F_(1,76)_=1.50, p>0.05, η^2^=0.019; F_(3,76)_=0.81, p>0.05, η^2^=0.031, respectively). Therefore, both sexes were analyzed together. Overall, all the diagnostic groups performed significantly worse in the eyes test than the control group. Thus, on average individuals with schizophrenia, schizoaffective and BPD received a lower score and make more mistakes in describing what the person in the picture was thinking or feeling compared to the healthy individuals (One way ANOVA, F_(3,83)_=9.835, p<0.001; Tukey’s, p<0.05; Figure 1A). No difference in the ability to decode the mental state of others was found between individuals with schizophrenia, schizoaffective, and BPD (Tukey’s, p>0.05).

**Figure 1:**
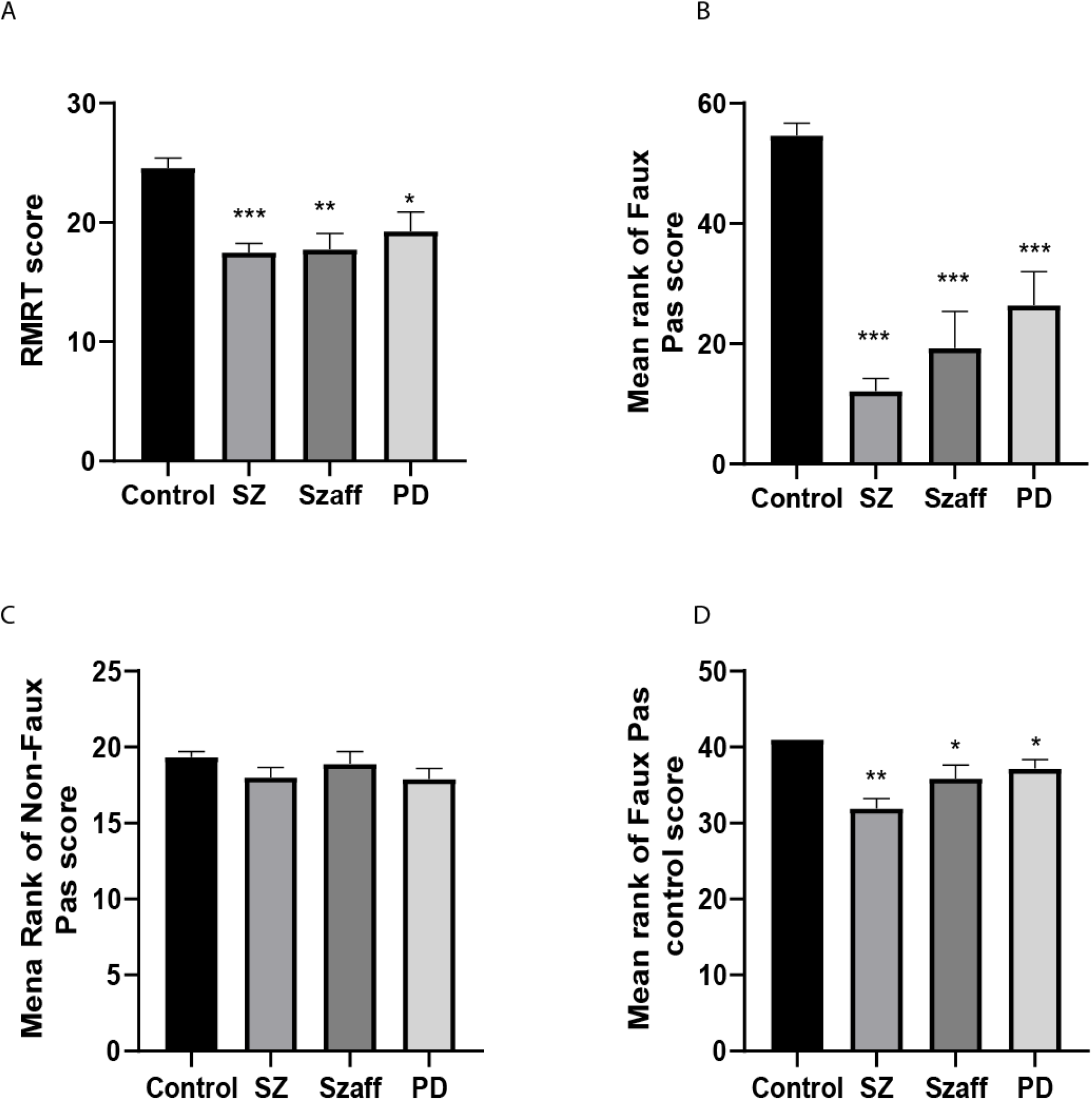
Individuals with schizophrenia, schizoaffective patients, and borderline personality disorder people are impaired in decoding the emotional state of others and in recognized Faux Pas motif. (A) Mean of the total eyes test score. B) Mean rank of recognized Faux Pas motif stories score B) Mean rank of recognized non-Faux Pas motif stories score. B) Mean rank of comprehension questions score. *\*p<0.05, **p<0.005, ***p<0.001* vs. control

### 3.3 Recognition of Faux Pas motif is impaired in individuals with schizophrenia, schizoaffective, and BPD

Analysis of the participant’s ability to recognize the Faux Pas motif reveals a significant main effect of diagnosis (F_(3,76)_=33.44, p<0.0001, η^2^=0.569), but not sex (F_(1,76)_=0.1.12, p>0.05, η ^2^ =0.15), without interaction between diagnosis and sex factors (F_(3,76)_=2.41, p>0.05, η ^2^ =0.087; S). Therefore, like in the analysis of the eyes test score, both sex groups were analyzed together. In comparison with the healthy individuals, all the three diagnosis groups recognized fewer Faux Pas motif stories (Kruskal-Wallis test, χ^2^_(3)_ =42.43, p<0.0001, N=84; adjusted using the Bonferroni correction, p<0.001). However, there was no significant difference between the three diagnosis groups (Bonferroni correction, p>0.05; Figure 2B). No difference was found between the three diagnostic groups and the control group in identifying the stories that did not contain the Faux Pax motif (Kruskal-Wallis test, χ^2^_(3)_ =4.48, p>0.05, N=84, Fig 2C). In comparison with the healthy individuals, all the three diagnosis groups received a lower score on the comprehension questions (Kruskal-Wallis test, χ^2^_(3)_ =31.19, p<0.0001, N=84; adjusted using the Bonferroni correction, p<0.001). However, there was no significant difference between the three diagnosis groups (Bonferroni correction, p>0.05).

### 3.4 Assessment of autistic symptoms and empathy

The degree of autistic symptoms and the level of empathy were assessed using the AQ and EQ questionnaires, respectively. Two-way ANOVA analysis revealed no significant main effect of sex effect in the AQ (F_(3,76)_=0.316, p>0.06, η^2^=0.004). Therefore, both sex groups were analyzed together. The mean scores and standard error of the mean of the AQ and EQ are presented in Table 2. Analysis of the results of the AQ questionnaire showed that individuals with schizophrenia and persons with BPD received a higher score on average in comparison with the control group (One-way ANOVA, F_(3,83)_=5.06, p<0.005; Tukey’s, p<0.05). No difference in the AQ scores was observed between individuals with schizoaffective and the control group (Tukey’s, p>0.05), and between the three diagnosis groups (Tukey’s, p>0.05). In contrast, analysis of the level of empathy showed a significant main effect of diagnosis and sex (F_(3,76)_=17.44, p<0.001, η^2^=0.408; F_(1,76)_=8.21, p<0.005, η^2^=0.098, respectively), with no interaction between the factors (F_(3,76)_=1.49, p>0.05, η^2^=0.056). Overall, the EQ scores were found to be lower in all the diagnostic groups as compared with the control group (Tukey’s, p<0.005). BPD individuals significantly scored higher than schizophrenic individuals did (Tukey’s, p<0.05). No difference in the EQ scores was observed between individuals with schizoaffective and the two-diagnostic groups: schizophrenia and BPD (Tukey’s, p>0.05;). Overall, the EQ scores were higher in the female group as compared with the male group (t_(82)_=-4.08, p<0.0001) and female PBD scored more than male PBD in the EQ (t_(9)_=-3.01, p<0.05). No difference in the EQ scores found between male and female schizophrenic, schizoaffective and control individuals (t_(42)_=-0.355, p>0.05; t_(9)_=-0.817, p>0.05; t_(16)_=-1.41, p>0.05, respectively)

**Table 2:** Mean score of AQ and EQ by group

| **Controls**<br>**(n=18)** | **Schizophrenia**<br>**(n=44)** | **Schizoaff**<br>**(n=11)** | **BPD**<br>**(n=11)** | **p-value<sup>a</sup>** |
| --- | --- | --- | --- | --- |
| **Autism-Spectrum Quotient** |  |  |  |  |
| 12.8±1.1 | 18.7.1±0.8 ** | 15.3±1.9 | 19.4±2.3 * | **p<0.005** |
| **Empathy quotient** |  |  |  |  |
| 56.3±2.3 | 33.9±1.5 ***<sup>#</sup> | 36.3±3.4 *** | 43.8±3.3 ** | **p<0.0001** |

Data presented as a mean ± standard error of the mean (SEM). <sup>a</sup>One-way ANOVA. \*p<0.05; \*\*p<0.001; \*\*\*p<0.0001 vs control. <sup>#</sup>p<0.05 vs BPD. Abbreviations: Schizoaff= Schizoaffective; BPD= Borderline Personality Disorder

### 3.5 Correlation analysis between the eyes test, Faux Pas, EQ, and AQ

Pearson’s correlation analyses showed that EQ scores were significantly correlated with the eyes test (R=0.599, R^2^=0.358, p<0.0001) and Faux Pas motif recognition (R=0.711, R^2^=0.505, p<0.0001). Furthermore, a significant correlation was found between the eyes test and Faux Pas motif recognition (R=0.697, R^2^=0.485, p<0.0001). No correlation was found between scores of AQ and the eyes test or Faux Pas motif recognition (R=-0.176, R^2^=0.030, p>0.05; R=-0.141, R^2^=0.019, p>0.05, respectively).

## 4. Discussion

Impairment of social abilities, interpersonal relationships, and social cognitive functioning characterize severe mental health and developmental disorders such as schizophrenia, depression, personality disorders, and autism. Studies have reported impairment in the ability to mentalize, understanding that others have their thoughts, beliefs, emotions, and intentions, among those diagnosed with schizophrenia, depression, and borderline personality disorder (Hegde et al., 2021; Maleki et al., 2020; Németh et al., 2018). There is no extended research to the best of our knowledge, which compared the level of mentalization between different mental health diagnoses. In the current study, we explored the mentalization capability of three groups of patients with different psychiatric diagnoses (schizophrenia, schizoaffective and borderline personality disorder), and evaluated their autistic symptoms and the level of empathy. The main outcome of the current pilot study is the ability to identify the emotional state of the others and recognize social motifs are impaired among all three diagnostic groups. Our preliminary results suggest that the cognitive and affective aspects of ToM are impaired among all the diagnostic groups. Additionally, we found a correlation between the degrees of empathy, but not autistic characteristics, and the level of performance in the mentalization tests, suggesting that the ability to infer the emotional experience of others might predict the level of the performance in ToM tasks.

Studies comparing the ToM ability of individuals with schizophrenia with schizoaffective patients are contradictory and include inconsistent results. Here, we demonstrated that the performance level of schizoaffective patients in both ToM tasks (decoding the emotion of the others and understanding an abnormal social situation) was similar and indistinguishable from individuals diagnosed with schizophrenia, and were significantly lower than the healthy individuals were. These results are in contradiction of our previous work conducted with a small group of schizoaffective patients (Tadmor et al., 2016), but support a recently published work (Hartman et al., 2019) and other works reported that the level of performance of ToM tasks was at the same extent (Hooper et al., 2010; Owoso et al., 2013; Pinna et al., 2014). In addition, the level of performance of these patients was also similar to the group of people diagnosed with borderline personality disorder. It is important to note that the level of the performance of BPD in the eyes test was also indistinguishable from the schizophrenic and schizoaffective individuals, but was different from the control group. The last result is going hand in hand with the published literature, which separately investigated BPD persons (Anupama et al., 2018; Németh et al., 2018; Richman and Unoka, 2015). A different pattern of social cognitive abilities was reported between women with schizophrenia and women with BPD. Although, schizophrenic women performed below women with BPD in a social cognition task the magnitude of the errors found to be higher in the BPD than in the schizophrenic group (Vaskinn et al., 2015). In another study, a different pattern of social cognition impairment between BPD and schizophrenia was reported. Schizophrenic individual’s error types were being under-mentalizing and those of BPD were over-mentalizing. Both individuals with schizophrenia and BPD underperformed the control group (Andreou et al., 2015). In the current study, we did not observe any difference in the errors pattern of mental state decoding and reasoning between schizophrenic, schizoaffective, and BPD individuals. However, individuals with schizophrenia made more errors in the recognition task and the comprehension questions than the other diagnostic groups, suggesting a more severe cognitive impairment of the schizophrenic subjects. Taken together, ToM performance is not associated with any of the three diagnostic groups per se, and highlight the need for more sensitive ToM tasks for future clinical diagnosis tool.

A correlation was found between the ability to mentalizing and empathy. Here, we report a positive correlation between the ability to identify the other’s emotions on the eyes test and the score of the empathy questionnaire. A person who understands and decodes the mental state of others probably will have higher empathy. This result is consistent with past studies which showed a correlation between the effective aspect of ToM and empathy (Hooker et al., 2008; Schnell et al., 2011). In addition, we also reported on a correlation between the EQ scores and recognition of social motifs. This result is not surprising, since it has been found that people with high empathy more successfully interpret and understand complex social events. For example, a correlation between empathy and the level of performance in false belief ToM tests was found (Ferguson et al., 2014). It is important to note that the control group and individuals with mental illness regardless of the diagnosis score above-average and average on the EQ questionnaire, respectively (Baron-Cohen and Wheelwright, 2004). These data suggest that on average, all the participants from the diagnosis groups have the ability for can understand how other people feel and respond appropriately. In contrast to the correlation found between ToM and empathy, no association was found between the cognitive and affective aspects of ToM and the AQ score. In addition, it is important to emphasize that the scores of all the research groups in the AQ questionnaire are in the normal range and do not indicate autistic-like traits or phenotype (Wheelwright et al., 2010). Together, results indicate that the impairment in the level of performance on both emotional and Faux Pas motif recognition tests probably did not result from autistic-related traits, as measured by AQ questionnaire, that characterizes schizophrenia and other psychiatric disorders.

To properly interpret the study findings, it is important to address its limitations. This initial study included a small number of schizoaffective and BPD patients and did not include recruitments from the outpatient clinics and the community. Although statistically significant differences were found, this study should be repeated in a larger group of subjects including, among others, not hospitalized patients. Second, in the BPD group, we also included individuals with comorbid affective disorders. Thus, we could not rule out that part of both the affective and cognitive impairment of ToM is simply due to the influences of this comorbidity. Finally, the duration of the illness of the schizophrenic and schizoaffective patients was longer than the BPD group, suggesting a chronic course of the illness probably with cognitive decline over the years that could explain why schizophrenic patients scoreless in the comprehension questions.

In summary, here we provide preliminary evidence that the cognitive and affective aspects of ToM are impaired in individuals with schizophrenia, schizoaffective, and BPD, and the level of the impairment did not differ between the diagnostic groups. Moreover, our data suggest that this impairment is associated with decreased empathy. Together, these data challenging the ability to use ToM as a differential diagnostic tool, and questioning the correlation between social skills and ToM performance.

## Data Availability

All data produced in the present study are available upon reasonable request to the authors

## Contributors

A.S designed this project. T.B, I.A, M.AL, and T.P performed the experiments. A.S and Y.A analyzed the data. A.S and T.B wrote the paper. I.G, E.P. and I.P. assisted with interpretation of the results. All the authors have contributed to and approved the final manuscript for publication.

## Acknowledgments

This study was supported by Mazor Mental Health Center, Akko, Israel (A.S, I.A and M. AA).

## Financial Disclosure

The authors report no financial relationship with any company or other commercial agents.

## Notes

### Competing Interest Statement

The authors have declared no competing interest.

### Author Declarations

The Manor Mental Health Center Helsinki committee approved the study (01-19-MZR).

